# Chest Pain Hospitalizations — How Emergency Clinician Tendencies Impact Care, Outcomes, and Costs

**DOI:** 10.64898/2026.10.02.26364626

**Authors:** Shih-Chuan Chou, Renee Y. Hsia, Fang Zhang, J. Frank Wharam

## Abstract

**Background:** The practice of hospitalizing emergency department (ED) patients with chest pain after ruling out acute myocardial infarction (AMI) varies widely. The long-term outcome and cost implications of this variation are uncertain.

**Methods:** Analyzing 2007–2021 claims from a national commercial insurer, we identified 223,569 adults (≥18 years) ED visits with a principal diagnosis of chest pain without major secondary cardiopulmonary diagnoses, seen by 19,001 clinicians in 2,273 EDs. Within each ED, clinicians with the highest and lowest terciles of risk-adjusted hospitalization rates were classified as high-versus low-admitting. Primary outcomes were 30-and 180-day subsequent AMI hospitalizations. Secondary outcomes included 7-day cardiac testing, 30-day coronary intervention, and 30-day total and out-of-pocket costs. We estimated adjusted rate ratios (aRRs) and relative cost differences using generalized estimating equations, adjusting for patient and visit characteristics.

**Results:** Patients had a mean age of 48.6 years, and 53.9% were female; measured characteristics were similar between groups. Compared with low-admitting clinicians, visits to high-admitting clinicians had higher 7-day cardiac testing (13.1% vs 10.8%; aRR 1.22, 95% CI 1.17–1.27) and 30-day coronary interventions (4.3% vs 3.8%; aRR 1.16, 95% CI 1.09–1.23). AMI hospitalization did not differ at 30 days (0.60% vs 0.60%; aRR 1.00, 95% CI 0.84–1.15) or 180 days (0.85% vs 0.87%; aRR 0.97, 95% CI 0.85–1.10). Thirty-day total costs were higher after visits to high-admitting clinicians (relative change 11.3%, 95% CI 8.8–13.8), and out-of-pocket costs ≥$1,000 were more common (32.0% vs 29.6%; relative change 8.2%, 95% CI 6.1–10.3).

**Conclusions:** Higher clinician hospitalizing tendency for ED chest pain is associated with greater downstream testing, more coronary interventions, and higher costs without lower short-or intermediate-term AMI risk. Reducing marginal admissions among high-admitting clinicians may decrease spending and practice variation with minimal impact on AMI outcomes.

## INTRODUCTION

Acute chest pain leads to over 7 million emergency department (ED) presentations annually.^1^ However, only approximately 5% of chest pain ED visits receive a diagnosis of acute coronary syndrome (ACS) or other significant cardiopulmonary condition.^2^ Hospitalizing chest pain patients in this scenario has been a common practice intended to minimize missed ACS^3^ and expedite coronary testing, with the goal of reducing the risk of downstream ACS or acute myocardial infarction (AMI). Although the American Heart Association (AHA) has historically endorsed the routine hospitalization and testing of patients presenting with chest pain,^4^ the revised guidelines of 2021 now emphasize care for patients classified as intermediate-or high-risk, and no longer advocate hospital admission for those categorized as low-risk.^5^ Today, chest pain remains one of the top reasons for short-stay hospitalizations,^6^ costing up to $3 billion annually,^7,8^ with additional unmeasured expenditures from downstream care cascades.^9,10^ Despite the substantial costs, the extent to which hospitalizations reduce the long-term risk of ACS and impose financial burdens on patients remains unclear.^11^

While a randomized trial including a “no hospitalization” arm could provide the most direct evidence, such a study is logistically and ethically challenging because hospitalization has become a standard practice. Recent studies have instead indirectly examined chest pain hospitalizations by comparing clinicians with varying admitting tendencies.^14,15^ These studies leveraged quasi-random assignment of patients to clinicians within the same ED, given that patients cannot select a specific clinician and the likelihood of seeing a particular clinician is not influenced by patient characteristics or preferences, thereby minimizing potential bias.^12^ Contrary to hospital-level comparisons,^13^ these studies have not found demonstrable differences in patient outcomes associated with clinicians’ hospitalization rates for chest pain patients.^14,15^ However, these studies did not assess cost implications or long-term outcomes. Moreover, these analyses were unable to capture subsequent care outside their systems because they were conducted within unique integrated health delivery systems (Veterans Affairs and Kaiser).

In this study, we built upon previous research to examine the associations between ED clinicians’ tendencies to hospitalize chest pain patients and subsequent care utilization, outcomes, and costs. We analyzed the claims database of a national commercial insurer, thereby extending prior research to encompass a nationwide population and thoroughly capturing each patient’s downstream care, including detailed out-of-pocket and total care costs. We hypothesized that ED visits for chest pain to clinicians with higher hospitalizing tendencies would be associated with increased downstream cardiac testing, invasive procedures, and higher costs, without significant alterations in long-term risk of AMI.

## METHODS

### Study Population

We drew our study population from approximately 60 million commercially or Medicare Advantage insured members within a de-identified claims database who were enrolled between January 1^st^, 2007 and September 30^th^, 2021. The dataset contains enrollment information for members and all medical, pharmacy, and hospitalization claims during enrollment periods, including ED visits, observation stays, and inpatient episodes. The study received exemption approval from the Institutional Review Board.

The study sample construction process is outlined in **Figure 1**, and detailed diagnosis and procedure code definitions are provided in **Supplemental Table S1**. We identified ED visits with a principal ED diagnosis of chest pain. We excluded visits with secondary diagnoses of AMI, pulmonary embolism, aortic disease, or other conditions that would account for chest pain symptoms and obviate further need for ACS evaluations. Additionally, we implemented a washout period, excluding chest pain ED visits if they were preceded within 6 months by any coronary testing, invasive interventions, or another chest pain ED visit, to focus on initial presentations rather than repeat evaluations.

**Figure 1.**
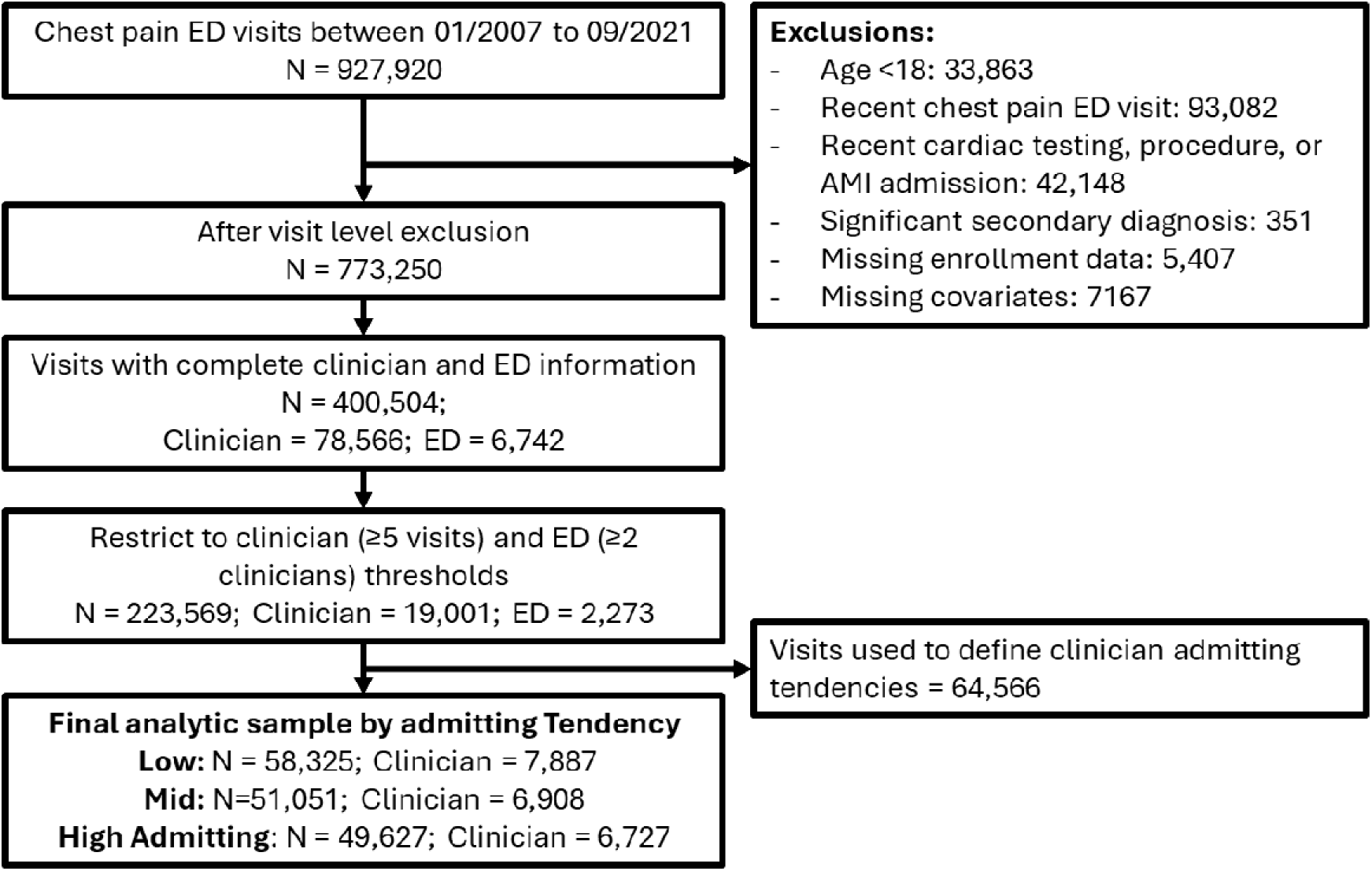
Study Sample Construction Flow Diagram **Abbreviation**: ED, emegency department; AMI, acute myocardial infarction **Note**: Recent ED visit and cardiac testing, procedures, or AMI admission is defined as within 6 months prior to an chest pain ED visit.

We excluded additional index chest pain ED visits of patients without an identifiable zip code (0.77%) or missing enrollment information (0.58%). Furthermore, we excluded patients younger than 18 years (3.65%) because the diagnostic considerations for pediatric and adult patients with chest pain differ substantially.

To facilitate within-ED between-clinician comparisons, we restricted our sample to visits with complete hospital and clinician identification. For clinicians who practiced at multiple EDs, we treated each clinician-ED combination as unique, recognizing that clinician behavior may vary across institutions due to differing practices and resources. Additionally, to establish clinician hospitalization tendencies based on prior ED visits, we further restricted our sample to ED visits managed by clinicians with at least 5 chest pain ED visits. Lastly, we excluded ED visits to hospitals with fewer than 2 included clinicians to allow within-ED comparisons.

### Visit Covariates

All modeling accounted for the following covariates, unless otherwise specified. We included available patient information such as age (in years) and sex assigned at birth. Additionally, we incorporated zip-code-based socioeconomic variables derived from 2014-2018 American Community Survey census tract data, including neighborhood poverty, education, and race. Furthermore, we included the zip code-level area deprivation index as a summary measure of socioeconomic status.^16^

To account for patient cardiovascular risk factors, we conducted a 1-year lookback from each index ED visit and identified diagnoses indicative of increased cardiovascular disease risk or general comorbidity (**Supplemental Table S1**). Additionally, we included calendar-year, month, and day-of-the-week fixed effects to account for secular, seasonal, and cyclical trends that may influence the outcomes.

### Study Outcomes

Our outcomes of interest included downstream care utilization, clinical outcomes, and costs. For care utilization, we first examined ***index hospitalizations***, defined as hospitalizations from the index chest pain ED visit, including both observation and inpatient hospitalizations. Additionally, we assessed downstream stress tests and non-invasive coronary imaging (**Supplemental Table S1**), within 7 and 30 days.^9,10^ Furthermore, we evaluated the occurrence of invasive angiography and revascularization procedures, including percutaneous coronary interventions (PCI) and coronary artery bypass grafting (CABG), within 30 and 180 days (**Supplemental Table S1**).

For clinical outcomes, we analyzed ***AMI hospitalizations***, defined as inpatient episodes with AMI listed as the first or second discharge diagnosis.^17^ Notably, hospital discharge diagnoses in our dataset were distinct from ED diagnoses. Inpatient or observation discharge diagnoses were derived from hospital care claims, while ED diagnoses were identified using ED-specific facility claims based on revenue codes (0450-9) and evaluation and management codes (99281-5). This distinction ensured that, for hospitalized ED visits, ED diagnoses reflected the clinical impression from the ED evaluation, rather than outcomes related to the subsequent hospitalization. We first analyzed ***index AMI hospitalizations***, defined as index hospitalizations with an AMI discharge diagnosis, representing cases where ischemia occurred or was identified during the hospital stay despite being absent in the ED evaluation. We also examined ***post-index AMI hospitalizations*** occurring within 7, 30, and 180 days after the index chest pain ED visit, excluding index AMI hospitalizations.

Lastly, we examined out-of-pocket and total standardized costs within 30 and 180 days to assess any associated differences or potential long-term savings. Specifically, we examined whether patient out-of-pocket costs reached $1,000 or $2,500 during the follow-up periods. All costs were standardized to 2020 dollars.

### Defining Clinician Hospitalizing Tendency

To establish clinician practice tendencies, we analyzed hospitalization (including both observation and inpatient stays) patterns from the first three chest pain ED visits per clinician (**Figure 2**).

**Figure 2.**
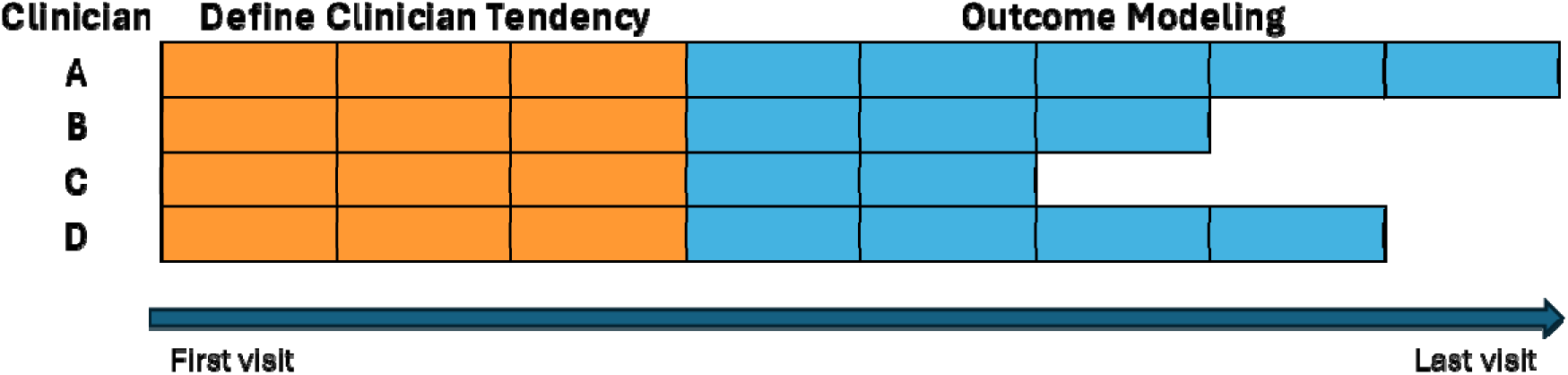
Study Design Illustration. **Note**. Orange cells represent first 3 visits used to model and calculate adjusted hospitalization tendencies for a specific clinician. Blue cells represent 4^th^ or later visits used for outcome modeling, with clinician hospitalizing tendency terciles as the primary variable.

We modeled the probability of hospitalization using ordinary least squares regression, controlling for the covariates described above. The model also included hospital (ED) fixed effects to account for institutional-level factors. We then calculated the clinician-level means of the regression residuals, representing each clinician’s deviation from the average clinician, given the same ED and patient visit. Using the mean clinician residuals, we then categorized clinicians into terciles within each ED, with top-tercile clinicians having the highest admitting tendencies. For EDs with only two clinician groups—such as having only two clinicians—clinicians were assigned to the top or bottom tercile, accordingly. There were no hospitals where all clinicians had identical mean residuals.

### Data Analysis

Our main analytic sample included all chest pain ED visits that were the fourth or subsequent visits for each clinician in each ED (**Figure 2**). This approach enabled us to use prior visits (first through third) as the basis for categorizing clinicians into terciles, which we then used to model outcomes from future visits (fourth and onwards). We assessed the validity of the quasi-randomization of patients by comparing patient characteristics and comorbid conditions across clinician hospitalizing tendency terciles and calculating standardized mean differences, with values <0.1 indicating minimal differences.^18^

To model the study outcomes, we used generalized estimating equations (GEE), accounting for clustering at the hospital level. The primary variable of interest was the clinician hospitalizing tendency terciles. The model included all patient and time covariates, as well as state fixed effects. For a more intuitive interpretation of the results, we used marginal estimating methods to calculate adjusted outcome rates for each clinician tercile and the adjusted rate ratios (aRR) compared to bottom-tercile clinicians.^19^

We conducted several prespecified sensitivity tests to assess the stability of our findings, with additional details in **Supplemental Methods**. First, we estimated reduced GEE models that included only the clinician hospitalizing tendency terciles and time variables. Second, we examined alternative definitions of clinicians’ hospitalizing tendencies, including a) using all index ED visits for each clinician (overall tendency) and b) based on all preceding index ED visits to the same clinician (cumulative tendency). Third, we examined a higher clinician inclusion threshold of 8 visits, defining clinician hospitalizing tendency using the first 5 visits.

We conducted several additional post-hoc subgroup analyses focusing on AMI hospitalization outcomes. We examined patients discharged or hospitalized during the index ED visits. We also examined patients by age (above and below 50 years) and by the presence of any comorbid conditions. Analyses were performed using SAS 9.4 (Cary, NC) and STATA 18 (College Station, TX).

## RESULTS

After exclusions and restricting to visit thresholds (**Figure 1**), we included 223,569 index ED visits with a principal diagnosis of chest pain. These visits represent care provided by 19,001 unique clinicians across 2,273 distinct EDs, totaling 21,524 unique ED-clinician combinations. To categorize clinicians in each ED into terciles of admitting tendency, we analyzed 64,566 visits, comprising the first three visits for all included clinicians. The final analytic sample for outcome modeling consisted of 159,003 index ED chest pain visits.

The overall patient sample had a mean age of 48.6 (standard deviation 17.6) years, with approximately 53.8% female (**Table 1 and Supplemental Table S2**). When comparing patients across clinician hospitalizing tendency terciles, patient characteristics, socioeconomic covariates, and comorbidities were well-balanced with no clinically meaningful differences.

**Table 1.** Patient Characteristics for Index Chest Pain ED Visits by Clinician Group.

| Clinician Admitting Tendency | Low |  | Mid |  | SMD | High |  | SMD |
| --- | --- | --- | --- | --- | --- | --- | --- | --- |
| n= | 58,325 |  | 51,051 |  |  | 49,627 |  |  |
| **Age, mean (SD)** | 48.8 | (17.6) | 48.9 | (17.9) | -0.002 | 49.8 | 17.9 | -0.053 |
| **Female, n(%)** | 31,518 | (54.0) | 27,445 | (53.8) | 0.006 | 26,784 | (54.0) | 0.001 |
| **Percent Neighborhood Demographics, mean (SD)** |  |  |  |  |  |  |  |  |
| Non-Hispanic White | 74.0 | (21.3) | 74.0 | (21.1) | 0.003 | 74.2 | 20.9 | -0.009 |
| Non-Hispanic Black | 14.3 | (19.4) | 14.4 | (19.4) | -0.005 | 14.2 | 19.1 | 0.005 |
| Hispanic | 15.0 | (17.2) | 15.0 | (17.2) | 0.000 | 15.1 | 17.2 | -0.009 |
| High School Education or Less | 36.8 | (14.9) | 36.8 | (14.9) | -0.004 | 37.1 | 14.8 | -0.026 |
| Income Below Poverty Level | 12.7 | (8.1) | 12.8 | (8.2) | -0.003 | 12.9 | 8.2 | -0.018 |
| **Neighborhood Income Below Poverty Level, N (%)** |  |  |  |  |  |  |  |  |
| <5.0% | 8,658 | (14.8) | 7,609 | (14.9) | 0.006 | 7,099 | (14.3) | 0.021 |
| 5.0%-9.9% | 18,213 | (31.2) | 15,852 | (31.1) |  | 15,322 | (30.9) |  |
| 10.0%-19.9% | 21,108 | (36.2) | 18,427 | (36.1) |  | 18,136 | (36.5) |  |
| ≥20.0% | 10,346 | (17.7) | 9,163 | (17.9) |  | 9,070 | (18.3) |  |
| **Neighborhood High School Education or Less, N (%)** |  |  |  |  |  |  |  |  |
| <15.0% | 4,376 | (7.5) | 3,670 | (7.2) | 0.012 | 3,374 | (6.8) | 0.030 |
| 15.0%-24.9% | 9,717 | (16.7) | 8,497 | (16.6) |  | 8,148 | (16.4) |  |
| 25.0%-39.9% | 19,702 | (33.8) | 17,367 | (34.0) |  | 16,793 | (33.8) |  |
| ≥40.0% | 24,530 | (42.1) | 21,517 | (42.1) |  | 21,312 | (42.9) |  |
| **Comorbidities, N(%)** |  |  |  |  |  |  |  |  |
| Coronary Artery Disease | 6,720 | (11.5) | 6,180 | (12.1) | 0.018 | 6,501 | (13.1) | 0.048 |
| Peripheral Vascular Disease | 1,611 | (2.8) | 1,445 | (2.8) | 0.004 | 1,521 | (3.1) | 0.018 |
| Congestive Heart Failure | 2,628 | (4.5) | 2,439 | (4.8) | 0.013 | 2,570 | (5.2) | 0.031 |
| Cerebral Vascular Disease | 2,157 | (3.7) | 1,964 | (3.8) | 0.008 | 2,018 | (4.1) | 0.019 |
| COPD | 2,802 | (4.8) | 2,570 | (5.0) | 0.011 | 2,555 | (5.1) | 0.016 |
| Chronic Kidney Disease | 2,749 | (4.7) | 2,497 | (4.9) | 0.008 | 2,567 | (5.2) | 0.021 |
| Diabetes | 6,981 | (12.0) | 6,428 | (12.6) | 0.019 | 6,861 | (13.8) | 0.055 |
| Hypertension | 16,908 | (29.0) | 15,055 | (29.5) | 0.011 | 15,355 | (30.9) | 0.043 |
| Hyperlipidemia | 16,455 | (28.2) | 14,552 | (28.5) | 0.006 | 14,736 | (29.7) | 0.033 |
| Obesity | 6,465 | (11.1) | 5,714 | (11.2) | 0.003 | 5,579 | (11.2) | 0.005 |
| Smoking History | 3,898 | (6.7) | 3,464 | (6.8) | 0.004 | 3,370 | (6.8) | 0.004 |
| **Area Deprivation Index, mean (SD)** | 47.1 | (22.5) | 47.3 | (22.5) | -0.006 | 48 | 22.4 | -0.037 |
| **Rural Residence, N (%)** | 6,040 | (10.4) | 4,763 | (9.3) | 0.034 | 5,133 | (10.3) | 0.000 |

**Abbreviation:** SMD, standardized mean difference; SD, standard deviation; COPD, chronic obstructive pulmonary disease.
**Note:** SMD reflects the comparison of 2nd or 3rd tercile with the 1st. SMD of <0.10 represents no meaningful difference.

**Table 2** compares downstream utilization associated with high-and low-admitting clinicians. Adjusted hospitalization rates were 18.0%, 20.8%, and 23.2% for the bottom-, middle-, and top-tercile clinicians, respectively. Top-and middle-tercile clinicians were associated with more downstream non-invasive cardiac testing within 7 days (aRR 1.22 [95%CI 1.17-1.27] and 1.10 [95%CI 1.06-1.14]) and 30 days (aRR 1.15 [95%CI 1.12-1.19] and 1.07 [95%CI 1.04-1.11]) after ED evaluations. Top and middle-tercile clinicians were also associated with higher rates of downstream coronary interventions within 30 days (aRR 1.16 [95%CI 1.09-1.23] and 1.09 [95%CI 1.03-1.16]) and 180 days (aRR 1.13 [95%CI 1.07-1.20] and 1.07 [95%CI 1.00-1.13]) of follow-up.

**Table 2.**
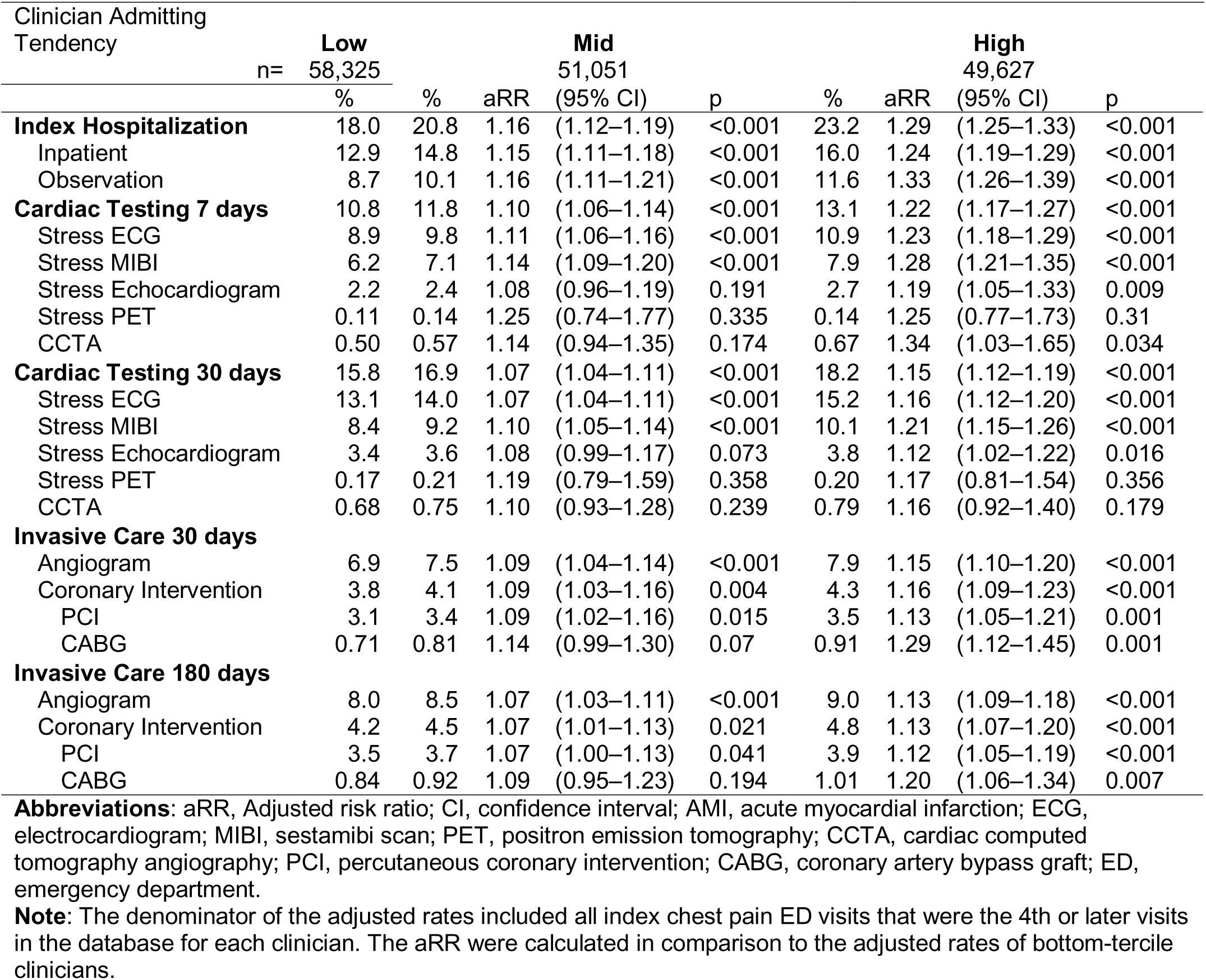
Adjusted Rates and Rate Ratios of Downstream Utilization by Clinician Group.

| Clinician Admitting<br>Tendency | Low<br>n= 58,325 |  | Mid<br>51,051 |  |  | High<br>49,627 |  |  |  |
| --- | --- | --- | --- | --- | --- | --- | --- | --- | --- |
|  | % | % | aRR | (95% CI) | p | % | aRR | (95% CI) | p |
| **Index Hospitalization** | 18.0 | 20.8 | 1.16 | (1.12–1.19) | <0.001 | 23.2 | 1.29 | (1.25–1.33) | <0.001 |
| Inpatient | 12.9 | 14.8 | 1.15 | (1.11–1.18) | <0.001 | 16.0 | 1.24 | (1.19–1.29) | <0.001 |
| Observation | 8.7 | 10.1 | 1.16 | (1.11–1.21) | <0.001 | 11.6 | 1.33 | (1.26–1.39) | <0.001 |
| **Cardiac Testing 7 days** | 10.8 | 11.8 | 1.10 | (1.06–1.14) | <0.001 | 13.1 | 1.22 | (1.17–1.27) | <0.001 |
| Stress ECG | 8.9 | 9.8 | 1.11 | (1.06–1.16) | <0.001 | 10.9 | 1.23 | (1.18–1.29) | <0.001 |
| Stress MIBI | 6.2 | 7.1 | 1.14 | (1.09–1.20) | <0.001 | 7.9 | 1.28 | (1.21–1.35) | <0.001 |
| Stress Echocardiogram | 2.2 | 2.4 | 1.08 | (0.96–1.19) | 0.191 | 2.7 | 1.19 | (1.05–1.33) | 0.009 |
| Stress PET | 0.11 | 0.14 | 1.25 | (0.74–1.77) | 0.335 | 0.14 | 1.25 | (0.77–1.73) | 0.31 |
| CCTA | 0.50 | 0.57 | 1.14 | (0.94–1.35) | 0.174 | 0.67 | 1.34 | (1.03–1.65) | 0.034 |
| **Cardiac Testing 30 days** | 15.8 | 16.9 | 1.07 | (1.04–1.11) | <0.001 | 18.2 | 1.15 | (1.12–1.19) | <0.001 |
| Stress ECG | 13.1 | 14.0 | 1.07 | (1.04–1.11) | <0.001 | 15.2 | 1.16 | (1.12–1.20) | <0.001 |
| Stress MIBI | 8.4 | 9.2 | 1.10 | (1.05–1.14) | <0.001 | 10.1 | 1.21 | (1.15–1.26) | <0.001 |
| Stress Echocardiogram | 3.4 | 3.6 | 1.08 | (0.99–1.17) | 0.073 | 3.8 | 1.12 | (1.02–1.22) | 0.016 |
| Stress PET | 0.17 | 0.21 | 1.19 | (0.79–1.59) | 0.358 | 0.20 | 1.17 | (0.81–1.54) | 0.356 |
| CCTA | 0.68 | 0.75 | 1.10 | (0.93–1.28) | 0.239 | 0.79 | 1.16 | (0.92–1.40) | 0.179 |
| **Invasive Care 30 days** |  |  |  |  |  |  |  |  |  |
| Angiogram | 6.9 | 7.5 | 1.09 | (1.04–1.14) | <0.001 | 7.9 | 1.15 | (1.10–1.20) | <0.001 |
| Coronary Intervention | 3.8 | 4.1 | 1.09 | (1.03–1.16) | 0.004 | 4.3 | 1.16 | (1.09–1.23) | <0.001 |
| PCI | 3.1 | 3.4 | 1.09 | (1.02–1.16) | 0.015 | 3.5 | 1.13 | (1.05–1.21) | 0.001 |
| CABG | 0.71 | 0.81 | 1.14 | (0.99–1.30) | 0.07 | 0.91 | 1.29 | (1.12–1.45) | 0.001 |
| **Invasive Care 180 days** |  |  |  |  |  |  |  |  |  |
| Angiogram | 8.0 | 8.5 | 1.07 | (1.03–1.11) | <0.001 | 9.0 | 1.13 | (1.09–1.18) | <0.001 |
| Coronary Intervention | 4.2 | 4.5 | 1.07 | (1.01–1.13) | 0.021 | 4.8 | 1.13 | (1.07–1.20) | <0.001 |
| PCI | 3.5 | 3.7 | 1.07 | (1.00–1.13) | 0.041 | 3.9 | 1.12 | (1.05–1.19) | <0.001 |
| CABG | 0.84 | 0.92 | 1.09 | (0.95–1.23) | 0.194 | 1.01 | 1.20 | (1.06–1.34) | 0.007 |

**Abbreviations:** aRR, Adjusted risk ratio; CI, confidence interval; AMI, acute myocardial infarction; ECG, electrocardiogram; MIBI, sestamibi scan; PET, positron emission tomography; CCTA, cardiac computed tomography angiography; PCI, percutaneous coronary intervention; CABG, coronary artery bypass graft; ED, emergency department.
**Note:** The denominator of the adjusted rates included all index chest pain ED visits that were the 4th or later visits in the database for each clinician. The aRR were calculated in comparison to the adjusted rates of bottom-tercile clinicians.

For AMI admissions, top-tercile clinicians was associated with higher rates of index AMI hospitalizations (aRR 1.12 [95%CI 1.04-1.20]; **Table 3**) compared to bottom-tercile clinicians. However, there were no significant differences in post-index AMI hospitalizations at 7 (aRR 1.00 [95%CI 0.82-1.18]), 30 (aRR 1.00 [95%CI 0.84-1.15]), and 180 days (aRR 0.97 [95%CI 0.85-1.10]). Comparison of middle-tercile clinicians with bottom-tercile clinicians showed similar patterns (**Table 3**).

**Table 3.** Rate of AMI Hospitalization by Clinician Groups.

| Clinician Admitting<br>Tendency |  | **Low** |  | **Mid** |  | **High** |  |  |
| --- | --- | --- | --- | --- | --- | --- | --- | --- |
|  | n= | 58,325 |  | 51,051 |  | 49,627 |  |  |
|  |  | % | % | aRR<br>(95% CI) | p | % | aRR<br>(95% CI) | p |
| **Index AMI** |  | 2.78 | 2.95 | 1.06<br>(0.98–1.14) | 0.118 | 3.12 | 1.12<br>(1.04–1.20) | 0.002 |
| **Post-Index AMI** |  |  |  |  |  |  |  |  |
| 7-day |  | 0.48 | 0.42 | 0.87<br>(0.72–1.02) | 0.091 | 0.48 | 1.00<br>(0.82–1.18) | 0.987 |
| 30-day |  | 0.60 | 0.53 | 0.87<br>(0.74–1.00) | 0.052 | 0.60 | 1.00<br>(0.84–1.15) | 0.952 |
| 180-day |  | 0.87 | 0.83 | 0.95<br>(0.83–1.07) | 0.376 | 0.85 | 0.97<br>(0.85–1.10) | 0.678 |

**Abbreviations:** AMI, acute myocardial infarction; aRR, adjusted risk ratio; CI, confidence interval
**Note:** The denominator of the adjusted rates included all index chest pain ED visits that were the 4th or later visits in the database for each clinician. The aRR were calculated in comparison to the adjusted rates of bottom-tercile clinicians.

Results were similar when we repeated our analysis with reduced outcome models (**Supplemental Table S3**), cumulative clinician hospitalizing tendency (**Supplemental Table S4**), and a higher visit threshold for clinician inclusion (**Supplemental Table S5**). However, when clinician hospitalizing tendencies were defined by all ED visits, top-tercile and middle-tercile clinicians were associated with significantly higher rates of post-index AMI hospitalizations (**Supplemental Table S6**).

In subgroup analyses by age and comorbidities (**Table 4**), we observed a similar pattern where higher clinician hospitalizing tendencies were associated with higher index AMI hospitalizations but no significant difference in post-index AMI hospitalizations.

**Table 4.** Adjusted AMI Hospitalization Rates by Clinician Groups in Patient Subgroups Clinician.

| Clinician<br>Admitting<br>Tendency | Low |  | Mid |  |  | High |  |  |  |
| --- | --- | --- | --- | --- | --- | --- | --- | --- | --- |
|  |  |  | INDEX VISIT DISPOSITION |  |  |  |  |  |  |
|  | % | % | aRR | (95% CI) | p | % | aRR | (95% CI) | p |
| **Discharged** | **n=47,482** | **n=39,956** |  |  |  | **n=36,854** |  |  |  |
| 7 day | 0.11 | 0.11 | 0.95 | (0.58–1.32) | 0.790 | 0.15 | 1.36 | (0.88–1.85) | 0.143 |
| 30 Days | 0.15 | 0.14 | 0.94 | (0.62–1.27) | 0.736 | 0.20 | 1.33 | (0.92–1.75) | 0.115 |
| 180 Days | 0.27 | 0.28 | 1.00 | (0.74–1.27) | 0.977 | 0.30 | 1.08 | (0.82–1.35) | 0.547 |
| **Admitted** | **n=10,843** | **n=11,095** |  |  |  | **n=12,773** |  |  |  |
| Index Visit | 14.4 | 13.5 | 0.94 | (0.88–1.01) | 0.087 | 13.4 | 0.93 | (0.87–0.99) | 0.029 |
| 7day | 2.0 | 1.6 | 0.77 | (0.63–0.92) | 0.002 | 1.6 | 0.79 | (0.63–0.94) | 0.007 |
| 30 Days | 2.5 | 1.9 | 0.77 | (0.64–0.91) | 0.001 | 2.0 | 0.80 | (0.66–0.94) | 0.004 |
| 180 Days | 3.3 | 2.8 | 0.85 | (0.72–0.98) | 0.019 | 2.7 | 0.83 | (0.71–0.95) | 0.005 |
|  |  |  | COMORBIDITIES |  |  |  |  |  |  |
|  | % | % | aRR | (95% CI) | p | % | aRR | (95% CI) | p |
| **No Comorbidities** | **n=31,907** | **n=27,742** |  |  |  | **n=26,236** |  |  |  |
| Index Visit | 0.98 | 1.20 | 1.22 | (1.03–1.42) | 0.024 | 1.29 | 1.32 | (1.10–1.54) | 0.004 |
| 7-Day | 0.25 | 0.18 | 0.70 | (0.45–0.95) | 0.018 | 0.23 | 0.89 | (0.58–1.20) | 0.488 |
| 30-Day | 0.27 | 0.21 | 0.78 | (0.52–1.04) | 0.091 | 0.25 | 0.92 | (0.62–1.23) | 0.627 |
| 180-Day | 0.33 | 0.29 | 0.88 | (0.62–1.14) | 0.356 | 0.31 | 0.92 | (0.65–1.20) | 0.581 |
| **With Comorbidities** | **n=26,418** | **n=23,309** |  |  |  | **n=23,391** |  |  |  |
| Index Visit | 5.0 | 5.1 | 1.02 | (0.94–1.10) | 0.602 | 5.3 | 1.07 | (0.99–1.16) | 0.092 |
| 7-Day | 0.8 | 0.7 | 0.96 | (0.77–1.14) | 0.652 | 0.8 | 1.06 | (0.84–1.27) | 0.606 |
| 30-days | 1.0 | 0.9 | 0.91 | (0.75–1.07) | 0.285 | 1.0 | 1.03 | (0.85–1.21) | 0.719 |
| 180-Days | 1.5 | 1.5 | 0.97 | (0.83–1.12) | 0.711 | 1.5 | 0.99 | (0.86–1.13) | 0.948 |
|  |  |  | AGE ABOVE BELOW 50 |  |  |  |  |  |  |
|  | % | % | aRR | (95% CI) | p | % | aRR | (95% CI) | p |
| **Below 50** | **n=31,604** | **n=27,570** |  |  |  | **n=25,794** |  |  |  |
| Index Visit | 0.47 | 0.57 | 1.21 | (0.93–1.48) | 0.140 | 0.65 | 1.39 | (1.07–1.70) | 0.016 |
| 7-Day | 0.14 | 0.11 | 0.77 | (0.42–1.12) | 0.198 | 0.13 | 0.93 | (0.48–1.37) | 0.748 |
| 30-Day | 0.17 | 0.12 | 0.74 | (0.42–1.05) | 0.104 | 0.15 | 0.91 | (0.50–1.32) | 0.663 |
| 180-Day | 0.19 | 0.15 | 0.76 | (0.46–1.06) | 0.111 | 0.18 | 0.93 | (0.56–1.30) | 0.720 |
| **Above 50** | **n=26,721** | **n=23,481** |  |  |  | **n=23,833** |  |  |  |
| Index | 5.5 | 5.7 | 1.04 | (0.96–1.12) | 0.310 | 6.0 | 1.09 | (1.01–1.17) | 0.024 |
| 7-Day | 0.9 | 0.8 | 0.89 | (0.72–1.06) | 0.200 | 0.9 | 1.01 | (0.81–1.21) | 0.933 |
| 30-Day | 1.1 | 1.0 | 0.89 | (0.74–1.04) | 0.155 | 1.1 | 1.01 | (0.83–1.18) | 0.928 |
| 180-Day | 1.7 | 1.6 | 0.97 | (0.84–1.11) | 0.679 | 1.6 | 0.98 | (0.84–1.11) | 0.745 |

**Abbreviations:** AMI, acute myocardial infarction; aRR, Adjusted risk ratio; CI, confidence interval
**Note:** The denominator of the adjusted rates included all index chest pain ED visits within each specific subgroup that were the 4th or later visits in the database for each clinician. The aRR were calculated in comparison to the adjusted rates of bottom-tercile clinicians.

When we examined only patients discharged from the index ED visits, the rates of post-index AMI hospitalizations at 7, 30, and 180 days were not significantly different across clinician terciles. When we examined hospitalized patients, top-tercile clinicians were associated with lower rates of index AMI admissions (aRR 0.93 [95%CI 0.87-0.99]) and post-index AMI admissions at 7, 30, and 180 days (aRR 0.79 [95%CI 0.63-0.94]; 0.80 [95%CI 0.66-0.94]; 0.83 [95%CI 0.71-0.95], respectively). We observed similar differences when we compared hospitalized patients between the middle-and bottom-tercile clinicians (**Table 4**).

ED visits managed by top-and middle-tercile clinicians resulted in 11.3% (95%CI 8.8-13.8) and 6.2% (95%CI 3.8-8.6) higher standardized care costs in 30-day periods (**Table 5**), compared to bottom-tercile clinicians. Out-of-pocket costs were also significantly higher, with top-and middle-tercile clinicians associated with 8.2% (95%CI 6.1-10.3) and 4.5% (95%CI 2.5-6.5) more patients responsible for costs of ≥$1,000 at 30 days, respectively.

**Table 5.**
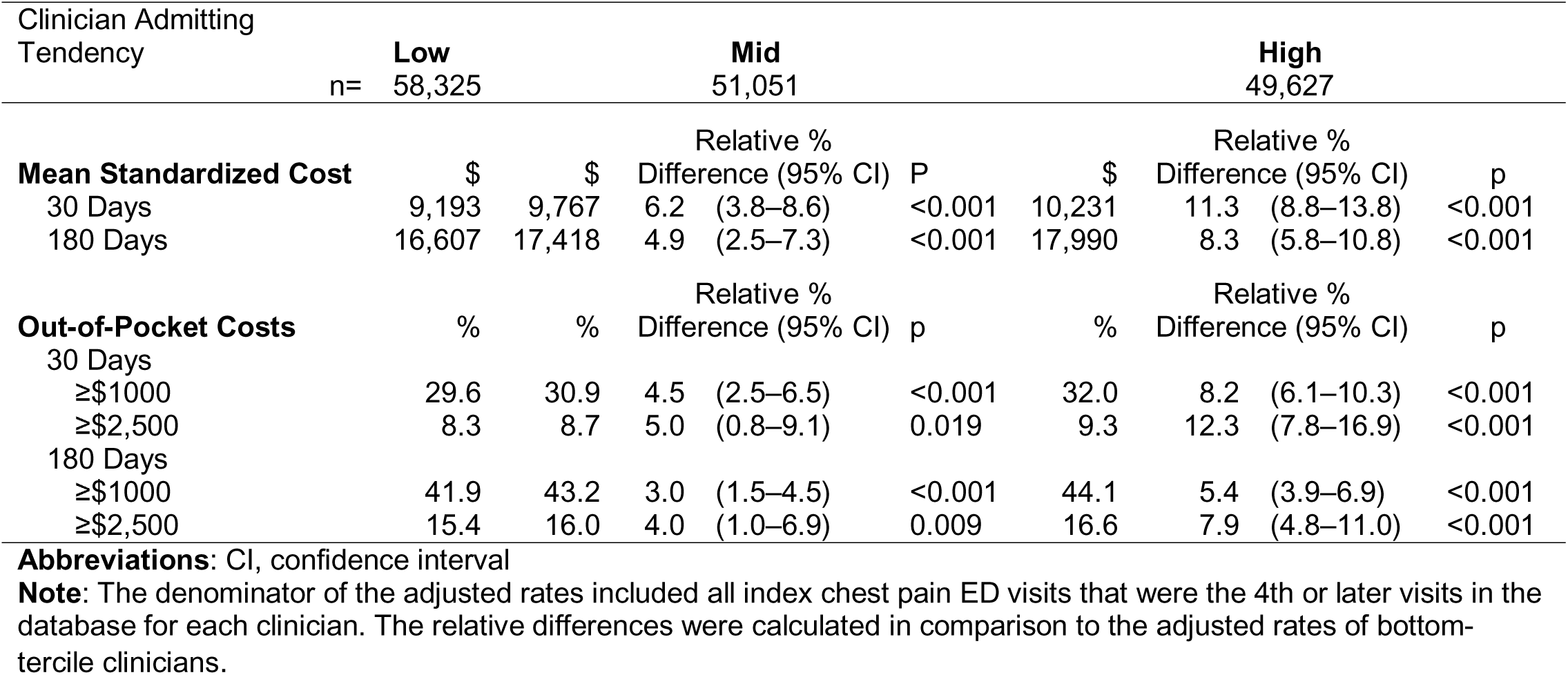
Adjusted Total Care and Out-of-Pocket Costs by Clinician Groups.

## DISCUSSION

In this study, we compared ED chest pain patients treated by clinicians with different hospitalizing tendencies. Overall, chest pain visits managed by clinicians with higher admitting tendencies were associated with up to 22% more downstream testing in 7 days and 16% more coronary interventions in 30 days compared to those managed by their low-admitting peers. The higher care utilization also significantly increased healthcare expenditures and patient financial burdens. However, despite the increased care intensity and costs, we observed no significant differences in long-term AMI risks.

After ruling out ACS and other significant cardiopulmonary conditions, one goal of hospitalizing ED patients with chest pain is to enable prompt evaluation for potential coronary atherosclerosis. Unsurprisingly, patients managed by high-admitting clinicians received more non-invasive tests and invasive coronary procedures. Most of these differences were present in the short-term, suggesting that, while ED clinicians primarily influence the decision to hospitalize and, potentially, pursue non-invasive testing^20,21^ during the index ED visits, these choices have a measurable influence on downstream care cascades.

Patients evaluated by high-admitting clinicians had 11.3% higher 30-day expenditures compared to low-admitting clinicians. This is especially notable considering hospitalizations for chest pain account for up to $3 billion annually in healthcare spending.^7,8^ Importantly, patients treated by high-admitting clinicians faced greater out-of-pocket costs, a financial burden over which they have little control, given that ED patients have limited influence on the type of clinician they will encounter. High admitting clinician practices can, therefore, expose additional patients to unexpected financial strain.^22,23^

Despite increased care utilization and costs, higher rates of chest pain hospitalizations did not lead to significant reductions in long-term AMI risks, even among older patients and those with cardiovascular comorbidities. These findings are consistent with prior studies in integrated health systems that showed no association between ED clinician practice variation and downstream cardiac outcomes.^14,15^

Notably, high-admitting clinician practices were associated with more AMI diagnoses during index hospitalizations. To better understand this, we conducted subgroup analyses of patients who were either discharged or hospitalized during their index ED visit. Among discharged patients, post-index AMI hospitalization rates were similar across clinician groups, indicating that lower-admitting clinicians were not more likely to release patients with clinically significant AMI. In contrast, among hospitalized patients, higher-admitting clinicians had lower rates of both index and post-index AMI hospitalizations, suggesting that these clinicians admitted lower-risk patients who might have been discharged by lower-admitting clinicians. Taken together—and given the minimal differences in patient characteristics across clinician groups—the additional AMI diagnoses observed during index hospitalizations among higher-admitting clinicians may have limited clinical relevance.

Our study leveraged the quasi-random distribution of patients to clinicians within each ED.^12^ Consistent with prior studies,^12,15,24^ we found comparable patient characteristics across clinician terciles without employing any matching or other balancing approaches, adding confidence that unobserved clinical or social factors were comparable across groups.

Our findings align with two prior clinician-level studies,^14,15^ but we improved upon their approach by utilizing prior visit patterns to predict future behavior. Earlier research classified clinicians’ hospitalizing tendencies based on all chest pain ED visits attributed to each clinician, introducing circular logic—where a clinician’s decision to hospitalize at a given visit could be influenced by both current and future behaviors, as well as past practices. This approach risks confounding by indication, as clinicians managing higher-risk patients, who were more likely to be hospitalized, would effectively be categorized as high-admitting clinicians.^25^ Sensitivity testing using this method (**Supplemental Table 6**) demonstrated the impact of this bias, underscoring the critical role of temporal factors in designing quasi-experimental observational studies. Although high clinician visit counts in prior studies may have mitigated this bias, our approach ensures a robust quasi-experimental design for settings with limited visits per clinician.

While we cannot directly assess the empirical value of chest pain hospitalizations, our study contributes to the growing evidence suggesting that marginal hospitalizations driven by different admitting thresholds among clinicians likely provide limited clinical benefit.^14,15,26^ Concurrently, it is important to recognize that most hospitalized patients would be admitted by any clinician. Advances in shared decision-making,^27^ clinical risk scores,^28–30^ and structured care pathways^31,32^ have been shown to reduce overall chest pain hospitalizations, but their ability to address clinician-level variation, especially among high-admitting providers, remains unclear. Clinician peer comparison, a strategy proven effective in areas such as opioid stewardship^33–35^ and imaging overuse,^36^ could complement these tools by benchmarking individual practices and “nudging” high-utilizing clinicians toward moderation. Future research should assess the feasibility and potential unintended consequences of this approach within emergency care settings.

Our study has several limitations. First, the insurance claims data we analyzed offered limited clinical granularity. However, the quasi-randomization of patients to clinicians enhances confidence that unobserved factors were plausibly balanced across the clinician terciles. Second, our dataset lacked mortality information, although AMI admissions remained a clinically relevant outcome for assessing the value of chest pain hospitalizations.^10^ Third, we excluded a considerable number of ED visits due to missing individual clinician or hospital identifiers, which likely stemmed from inconsistent billing practices. We also excluded many ED clinicians below the visit threshold. As such, our findings may generalize to ED clinicians who see a moderate number of commercially insured patients.

## CONCLUSION

In this analysis of ED visits for chest pain, we found that higher clinician hospitalization rates were associated with increased downstream coronary testing and invasive procedures. This increased expenditure and out-of-pocket cost burden among patients, yet did not reduce longer-term AMI risk. Future studies should examine potential drivers of clinician practice variation, including the impact of risk stratification tools. Our findings support interventions to reduce marginal hospitalizations by high-admitting clinicians, thereby decreasing unnecessary downstream care cascades and associated costs.

## Data Availability

Data sharing is prohibited by the data use agreement with the data vendor. All analytic codes are available upon request to the authors.

## ACKNOWLEDGEMENT

None

## SUPPLEMENTAL MATERIAL

Supplemental Methods

Tables S1–S6

