## Supplemental Tables for "Chest Pain Hospitalizations — How Emergency Clinician Tendencies Impact Care, Outcomes, and Costs"

**SUPPLEMENTAL MATERIALS**

**Supplemental Methods**

**Table S1. Definitions for diagnoses and procedures**

**Table S2. Time for Chest Pain ED Visits by Clinician Group**

**Table S3. Adjusted Rates of Downstream Utilization and AMI Hospitalization by Clinician Groups, Reduced Model**

**Table S4. Adjusted Rates of Downstream Utilization and AMI Hospitalization by Clinician Cumulative Tendency**

**Table S5. Adjusted Rates of Downstream Utilization and AMI Hospitalization by Clinician Hospitalizing Tendency, among Clinicians with ≥8 Visits.**

**Table S6. Adjusted Rates of Downstream Utilization and AMI Hospitalization by Clinician Hospitalizing Tendency, Defined by All ED Visits.**

**Supplemental Methods.**

I. **Clinician admitting tendency based on all ED visits. (Overall Tendency)**

In this sensitivity test, instead of the first 3 visits, we used ***all included index chest pain ED visits*** to define clinician hospitalizing tendencies. We applied the same ordinary least squares (OLS) model described in the main article and calculated the regression residuals for each visit. For each clinician, we then calculated the mean residual across all their index ED visits. We similarly categorized clinicians into terciles based on comparisons of their mean residuals with those of other clinicians within the same ED, and modeled outcomes using generalized estimating equations (GEEs) with clinician tercile as the primary predictor.

II. **Clinician admitting tendency based on preceding ED visits. (Cumulative Tendency)**

Similar to above, we again included all index ED visits and calculated regression residuals for each visit using the OLS model. However, instead of calculating clinician-level mean residual across all visits, we calculated a cumulative mean residual for each visit after a clinician’s first three visits. For each clinician's 4^th^ and subsequent visits, the cumulative mean residual for any given visit was derived by averaging the residuals from the clinician’s first three visits and all prior visits up to the visit in question. For instance, the cumulative mean residual for a clinician’s 5^th^ visit was calculated using the residuals from their 1^st^ through 4^th^ visits. This method allowed us to account for potential changes in a clinician’s hospitalizing tendencies over time and does not suffer from the potential bias from using future visits to predict current behavior. We categorized ***visits*** (rather than clinicians) into terciles of cumulative mean residuals within each ED. Outcomes were then modeled using the same GEE framework, with admitting tercile as the primary predictor variable.

**III. Clinician inclusion threshold of eight visits and admitting tendency defined using the first five visits.**

In this sensitivity analysis, we raised the clinician inclusion threshold to require at least 8 index chest pain ED visits, excluding visits to EDs with fewer than 2 clinicians that meet the eight-visit threshold. We then used the first 5 visits of each clinician to characterize their practice and categorize terciles of admitting tendencies using a similar OLS model outlined in the main methods. While this higher threshold resulted in a smaller sample of chest pain ED visits, clinician attribution into terciles may be more accurate. We then modeled study outcomes as a function of clinician terciles as the primary predictor variable and other covariates, using similar GEE models but with the 6^th^ and subsequent visits to each clinician.

| **Table S1. Definitions for diagnoses and procedures** | |
| --- | --- |
| **Diagnoses** | **CPT or ICD codes** |
| Chest pain | 786.50, 786.51, 786.59; R07.1, R07.2, R07.89, and R07.9 |
| ***Chest pain diagnoses for exclusion*** |  |
| Acute myocardial infarction | 410.xx; I21.xx |
| Acute or subacute ischemic heart disease (unstable angina) | 411.1, 411.8x; I22.xx, I23.xx, I24.xx, I25.xx |
| Angina | 413; I20.xx |
| Acute pulmonary heart disease (pulmonary embolism) | 415.xx; I26.xx |
| Aortic aneurysm and dissection | 441.xx; I71.0x, I71.1x, I71.3x, I71.5x, I71.8x |
| ***Non-invasive procedures*** |  |
| Stress electrocardiogram | 93015-93018 |
| Myocardial perfusion scan | 78451-78454 |
| Stress echocardiogram | 93350, 93351 |
| Stress positron emission testing | 78491-2, 78430-1 |
| Coronary computed tomography angiogram | 75574 |
| ***Invasive procedures*** |  |
| Coronary angiography (catheterization) | 93452-93462, 93508, 93510, 93526, 93539, 93540, 93543, 93545. |
| Percutaneous coronary intervention | 92920-92944, 92973-5, 92980-92982, 92984, 92995, 92996, G0290, G0291, C9600-8). |
| Coronary artery bypass grafting | 33510-33536 |
| ***Cardiovascular risk factors and comorbidities*** |  |
| Coronary artery disease | 410.xx-414.xx; I20.xx-I25.xx |
| Peripheral vascular disease | 440.xx; I70.xx |
| Congestive heart failure | 428.xx; I50.xx |
| Cerebral vascular disease | 433.xx-436.xx; I63x-I66x |
| Chronic obstructive pulmonary disease | 491.xx, 492.xx; J41.0x, J42.xx, J43.x, J44.x |
| Chronic Kidney Disease | 585.xx; N18x |
| Diabetes | 249.xx, 250.xx; E08.xx-E11.xx, E13.xx |
| Hypertension | 401.xx-405.xx; I10.xx-I15.xx |
| Dyslipidemia | 272.xx; E78.xx |
| Obesity | 278.0x; E66.xx |
| Smoking | 305.1x; F17.21x |
| **Abbreviations**: CPT, common procedure terminology; ICD, international classification of diseases | |

| **Table S2. Time of Chest Pain ED Visits by Clinician Group** | | | | | | | | |
| --- | --- | --- | --- | --- | --- | --- | --- | --- |
| Clinician Group | Low Admitting | | Mid | |  | High | |  |
| n= | 58,325 | | 51,051 | | SMD | 49,627 | | SMD |
| **Day of Week, N (%)** |  |  |  |  |  |  |  |  |
| Sunday | 6987 | (12.0) | 6110 | (12.0) | 0.016 | 6017 | (12.1) | 0.02 |
| Monday | 10078 | (17.3) | 8724 | (17.1) |  | 8391 | (16.9) |  |
| Tuesday | 9136 | (15.7) | 7830 | (15.3) |  | 7510 | (15.1) |  |
| Wednesday | 8648 | (14.8) | 7508 | (14.7) |  | 7442 | (15.0) |  |
| Thursday | 8521 | (14.6) | 7467 | (14.6) |  | 7357 | (14.8) |  |
| Friday | 8172 | (14.0) | 7334 | (14.4) |  | 7035 | (14.2) |  |
| Saturday | 6783 | (11.6) | 6078 | (11.9) |  | 5875 | (11.8) |  |
| **Calendar Month, N (%)** | |  |  |  |  |  |  |  |
| 1 | 4963 | (8.5) | 4359 | (8.5) | 0.029 | 4284 | (8.6) | 0.019 |
| 2 | 4895 | (8.4) | 4328 | (8.5) |  | 4174 | (8.4) |  |
| 3 | 5354 | (9.2) | 4602 | (9.0) |  | 4416 | (8.9) |  |
| 4 | 4857 | (8.3) | 4274 | (8.4) |  | 4135 | (8.3) |  |
| 5 | 5068 | (8.7) | 4323 | (8.5) |  | 4294 | (8.7) |  |
| 6 | 4894 | (8.4) | 4203 | (8.2) |  | 4093 | (8.2) |  |
| 7 | 5064 | (8.7) | 4355 | (8.5) |  | 4402 | (8.9) |  |
| 8 | 4946 | (8.5) | 4367 | (8.6) |  | 4234 | (8.5) |  |
| 9 | 4726 | (8.1) | 4504 | (8.8) |  | 4194 | (8.5) |  |
| 10 | 4572 | (7.8) | 3927 | (7.7) |  | 3847 | (7.8) |  |
| 11 | 4433 | (7.6) | 3824 | (7.5) |  | 3700 | (7.5) |  |
| 12 | 4553 | (7.8) | 3985 | (7.8) |  | 3854 | (7.8) |  |
| **Year, N (%)** |  |  |  |  |  |  |  |  |
| 2007 | 577 | (1.0) | 510 | (1.0) | 0.097 | 658 | (1.3) | 0.072 |
| 2008 | 1328 | (2.3) | 1012 | (2.0) |  | 1325 | (2.7) |  |
| 2009 | 1690 | (2.9) | 1267 | (2.5) |  | 1643 | (3.3) |  |
| 2010 | 1914 | (3.3) | 1498 | (2.9) |  | 1807 | (3.6) |  |
| 2011 | 2564 | (4.4) | 1794 | (3.5) |  | 2232 | (4.5) |  |
| 2012 | 3521 | (6.0) | 2643 | (5.2) |  | 2901 | (5.8) |  |
| 2013 | 3581 | (6.1) | 2797 | (5.5) |  | 3010 | (6.1) |  |
| 2014 | 3453 | (5.9) | 2812 | (5.5) |  | 2755 | (5.6) |  |
| 2015 | 4318 | (7.4) | 3727 | (7.3) |  | 3215 | (6.5) |  |
| 2016 | 6190 | (10.6) | 5403 | (10.6) |  | 4902 | (9.9) |  |
| 2017 | 6436 | (11.0) | 5859 | (11.5) |  | 5366 | (10.8) |  |
| 2018 | 7052 | (12.1) | 6653 | (13.0) |  | 6020 | (12.1) |  |
| 2019 | 6768 | (11.6) | 6227 | (12.2) |  | 5740 | (11.6) |  |
| 2020 | 4998 | (8.6) | 4926 | (9.6) |  | 4552 | (9.2) |  |
| 2021 | 3935 | (6.7) | 3923 | (7.7) |  | 3501 | (7.1) |  |
| **Abbreviation**: SMD, standardized mean difference | | | | | | | | |

| **Table S3. Adjusted Rates of Downstream Utilization and AMI Hospitalization by Clinician Groups, Reduced Model** | | | | | | | | | |
| --- | --- | --- | --- | --- | --- | --- | --- | --- | --- |
| Clinician Admitting Tendency | **Low** | **Mid** | | | | **High** | | | |
| n= | 58,325 | 51,051 | | | | 49,627 | | | |
|  | % | % | aRR | (95% CI) | p | % | aRR | (95% CI) | p |
| **Index Hospitalization** | 18.4 | 21.1 | 1.14 | (1.11–1.18) | <0.001 | 24.4 | 1.32 | (1.28–1.37) | <0.001 |
| Inpatient | 13.3 | 15.1 | 1.14 | (1.09–1.18) | <0.001 | 17.1 | 1.28 | (1.23–1.34) | <0.001 |
| Observation | 8.8 | 10.1 | 1.15 | (1.09–1.20) | <0.001 | 12.0 | 1.36 | (1.29–1.43) | <0.001 |
| **Cardiac Testing 7 days** | 10.8 | 11.7 | 1.09 | (1.04–1.13) | <0.001 | 13.3 | 1.24 | (1.19–1.29) | <0.001 |
| Stress ECG | 8.9 | 9.8 | 1.09 | (1.04–1.15) | <0.001 | 11.2 | 1.25 | (1.19–1.31) | <0.001 |
| Stress MIBI | 6.2 | 7.1 | 1.13 | (1.07–1.19) | <0.001 | 8.2 | 1.31 | (1.24–1.39) | <0.001 |
| Stress Echocardiogram | 2.2 | 2.3 | 1.06 | (0.95–1.18) | 0.295 | 2.6 | 1.19 | (1.05–1.34) | 0.010 |
| Stress PET | 0.11 | 0.14 | 1.25 | (0.77–1.77) | 0.353 | 0.15 | 1.32 | (0.81–1.83) | 0.223 |
| CCTA | 0.50 | 0.56 | 1.13 | (0.92–1.34) | 0.237 | 0.66 | 1.34 | (1.02–1.65) | 0.036 |
| **Cardiac Testing 30 days** | 15.7 | 16.7 | 1.06 | (1.03–1.10) | <0.001 | 18.3 | 1.17 | (1.13–1.20) | <0.001 |
| Stress ECG | 13.1 | 13.9 | 1.06 | (1.03–1.10) | <0.001 | 15.4 | 1.17 | (1.13–1.21) | <0.001 |
| Stress MIBI | 8.5 | 9.2 | 1.08 | (1.04–1.13) | <0.001 | 10.4 | 1.23 | (1.18–1.29) | <0.001 |
| Stress Echocardiogram | 3.3 | 3.5 | 1.07 | (0.98–1.16) | 0.137 | 3.7 | 1.12 | (1.02–1.23) | 0.021 |
| Stress PET | 0.17 | 0.20 | 1.18 | (0.77–1.58) | 0.392 | 0.21 | 1.23 | (0.85–1.60) | 0.239 |
| CCTA | 0.66 | 0.72 | 1.09 | (0.91–1.27) | 0.320 | 0.77 | 1.17 | (0.92–1.42) | 0.184 |
| **Invasive Care 30 days** |  |  |  |  |  |  |  |  |  |
| Angiogram | 6.9 | 7.5 | 1.09 | (1.04–1.13) | <0.001 | 8.3 | 1.20 | (1.14–1.25) | <0.001 |
| Coronary Intervention | 3.7 | 4.1 | 1.10 | (1.03–1.16) | 0.007 | 4.5 | 1.21 | (1.14–1.29) | <0.001 |
| PCI | 3.1 | 3.3 | 1.09 | (1.01–1.17) | 0.019 | 3.6 | 1.19 | (1.10–1.27) | <0.001 |
| CABG | 0.69 | 0.80 | 1.15 | (0.99–1.31) | 0.062 | 0.94 | 1.36 | (1.19–1.53) | <0.001 |
| **Invasive Care 180 days** |  |  |  |  |  |  |  |  |  |
| Angiogram | 8.0 | 8.6 | 1.07 | (1.02–1.11) | 0.004 | 9.5 | 1.18 | (1.13–1.23) | <0.001 |
| Coronary Intervention | 4.2 | 4.5 | 1.07 | (1.01–1.13) | 0.029 | 5.0 | 1.19 | (1.12–1.26) | <0.001 |
| PCI | 3.4 | 3.7 | 1.07 | (1.00–1.14) | 0.050 | 4.0 | 1.17 | (1.10–1.25) | <0.001 |
| CABG | 0.83 | 0.91 | 1.10 | (0.96–1.25) | 0.172 | 1.05 | 1.27 | (1.11–1.42) | <0.001 |
| **AMI Admissions** |  |  |  |  |  |  |  |  |  |
| Index | 2.7 | 2.9 | 1.06 | (0.98–1.14) | 0.125 | 3.2 | 1.18 | (1.09–1.26) | <0.001 |
| 7 day | 0.48 | 0.42 | 0.87 | (0.72–1.02) | 0.081 | 0.50 | 1.04 | (0.85–1.22) | 0.693 |
| 30 day | 0.60 | 0.52 | 0.87 | (0.73–1.00) | 0.049 | 0.62 | 1.04 | (0.88–1.20) | 0.626 |
| 180 days | 0.86 | 0.81 | 0.95 | (0.83–1.07) | 0.407 | 0.89 | 1.03 | (0.90–1.16) | 0.660 |
| **Abbreviations**: aRR, Adjusted risk ratio; CI, confidence interval; AMI, acute myocardial infarction; ECG, electrocardiogram; MIBI, sestamibi scan; PET, positron emission tomography; CCTA, cardiac computed tomography angiography; PCI, percutaneous coronary intervention; CABG, coronary artery bypass graft; ED, emergency department. | | | | | | | | | |
| **Note**: The denominator of the adjusted rates included all index chest pain ED visits that were the 4^th^ or later visits in the database for each clinician. Adjusted rates were calculated using generalized estimating equations that included only time variables and no other covariates, similarly accounting for clustering at the hospital level. The aRR were calculated in comparison to the adjusted rates of bottom-tercile clinicians. | | | | | | | | | |

| **Table S4. Adjusted Rates of Downstream Utilization and AMI Hospitalization by Clinician Cumulative Tendency** | | | | | | | | | | | | | |
| --- | --- | --- | --- | --- | --- | --- | --- | --- | --- | --- | --- | --- | --- |
| Clinician Cumulative Admitting Tendency | **Low** | | **Mid** | | | | | | | **High** | | | |
| n= | 53,786 | | 52,973 | | | | | | | 52,244 | | | |
|  | **%** | | **%** | | **aRR** | | **(95% CI)** | | p | **%** | **aRR** | **(95% CI)** | p |
| **Hospitalization** | 17.4 | | 20.3 | | 1.16 | | (1.13–1.19) | | <0.001 | 24.1 | 1.38 | (1.34–1.43) | <0.001 |
| Inpatient | 12.4 | | 14.5 | | 1.18 | | (1.14–1.21) | | <0.001 | 16.7 | 1.35 | (1.30–1.41) | <0.001 |
| Observation | 8.7 | | 9.7 | | 1.12 | | (1.08–1.17) | | <0.001 | 11.8 | 1.36 | (1.29–1.43) | <0.001 |
| **Cardiac Testing 7 days** | 10.3 | | 11.6 | | 1.13 | | (1.08–1.17) | | <0.001 | 13.6 | 1.32 | (1.26–1.38) | <0.001 |
| Stress ECG | 8.6 | | 9.7 | | 1.14 | | (1.09–1.18) | | <0.001 | 11.2 | 1.31 | (1.24–1.37) | <0.001 |
| Stress MIBI | 5.9 | | 6.9 | | 1.18 | | (1.12–1.23) | | <0.001 | 8.2 | 1.40 | (1.32–1.47) | <0.001 |
| Stress Echocardiogram | 2.2 | | 2.4 | | 1.09 | | (0.99–1.20) | | 0.077 | 2.7 | 1.24 | (1.09–1.39) | 0.002 |
| Stress PET | 0.12 | | 0.14 | | 1.14 | | (0.79–1.50) | | 0.43 | 0.13 | 1.11 | (0.64–1.59) | 0.635 |
| CCTA | 0.46 | | 0.52 | | 1.14 | | (0.90–1.38) | | 0.244 | 0.76 | 1.67 | (1.23–2.11) | 0.003 |
| **Cardiac Testing 30 days** | 15.4 | | 16.7 | | 1.09 | | (1.05–1.12) | | <0.001 | 18.6 | 1.21 | (1.17–1.25) | <0.001 |
| Stress ECG | 12.8 | | 14.0 | | 1.10 | | (1.06–1.14) | | <0.001 | 15.4 | 1.21 | (1.16–1.25) | <0.001 |
| Stress MIBI | 8.1 | | 9.1 | | 1.12 | | (1.08–1.17) | | <0.001 | 10.3 | 1.27 | (1.21–1.33) | <0.001 |
| Stress Echocardiogram | 3.4 | | 3.6 | | 1.06 | | (0.98–1.14) | | 0.166 | 3.8 | 1.12 | (1.02–1.22) | 0.015 |
| Stress PET | 0.18 | | 0.20 | | 1.09 | | (0.79–1.39) | | 0.573 | 0.20 | 1.10 | (0.74–1.45) | 0.596 |
| CCTA | 0.62 | | 0.68 | | 1.10 | | (0.90–1.29) | | 0.324 | 0.92 | 1.50 | (1.16–1.83) | 0.003 |
| **Invasive Care 30 days** |  | |  | |  | |  | |  |  |  |  |  |
| Angiogram | 6.7 | | 7.4 | | 1.10 | | (1.05–1.15) | | <0.001 | 8.1 | 1.20 | (1.15–1.26) | <0.001 |
| Coronary Intervention | 3.7 | | 4.1 | | 1.13 | | (1.06–1.20) | | <0.001 | 4.4 | 1.19 | (1.12–1.26) | <0.001 |
| PCI | 3.0 | | 3.4 | | 1.12 | | (1.05–1.20) | | <0.001 | 3.6 | 1.17 | (1.09–1.25) | <0.001 |
| CABG | 0.71 | | 0.80 | | 1.13 | | (0.99–1.28) | | 0.076 | 0.90 | 1.27 | (1.09–1.44) | 0.003 |
| **Invasive Care 180 days** |  | |  | |  | |  | |  |  |  |  |  |
| Angiogram | 7.8 | | 8.4 | | 1.08 | | (1.03–1.12) | | <0.001 | 9.2 | 1.18 | (1.13–1.23) | <0.001 |
| Coronary Intervention | 4.1 | | 4.5 | | 1.11 | | (1.05–1.17) | | <0.001 | 4.8 | 1.16 | (1.10–1.23) | <0.001 |
| PCI | 3.4 | | 3.7 | | 1.10 | | (1.03–1.17) | | 0.004 | 3.9 | 1.16 | (1.09–1.23) | <0.001 |
| CABG | 0.84 | | 0.93 | | 1.10 | | (0.97–1.24) | | 0.136 | 1.00 | 1.18 | (1.03–1.33) | 0.016 |
| **AMI Admissions** | |  | |  | |  | |  | |  |  |  |  |
| Index | 2.7 | | 3.0 | | 1.11 | | (1.03–1.19) | | 0.007 | 3.1 | 1.13 | (1.05–1.21) | 0.001 |
| 7 day | 0.47 | | 0.44 | | 0.94 | | (0.77–1.11) | | 0.475 | 0.47 | 1.00 | (0.83–1.17) | 0.979 |
| 30 day | 0.59 | | 0.56 | | 0.95 | | (0.80–1.11) | | 0.561 | 0.58 | 0.99 | (0.84–1.14) | 0.919 |
| 180 days | 0.88 | | 0.83 | | 0.94 | | (0.82–1.07) | | 0.374 | 0.84 | 0.96 | (0.83–1.08) | 0.485 |
| **Abbreviations**: aRR, Adjusted risk ratio; CI, confidence interval; AMI, acute myocardial infarction; ECG, electrocardiogram; MIBI, sestamibi scan; PET, positron emission tomography; CCTA, cardiac computed tomography angiography; PCI, percutaneous coronary intervention; CABG, coronary artery bypass graft; ED, emergency department. | | | | | | | | | | | | | |
| **Note**: The denominator of the adjusted rates included all index chest pain ED visits that were the 4^th^ or later visits in the database for each clinician. The aRR were calculated in comparison to the adjusted rates of bottom-tercile clinicians. | | | | | | | | | | | | | |

| **Table S5. Adjusted Rates of Downstream Utilization and AMI Hospitalization by Clinician Admitting Tendency, among Clinicians with ≥8 Visits.** | | | | | | | | | |
| --- | --- | --- | --- | --- | --- | --- | --- | --- | --- |
| Clinician Admitting Tendency | **Low** | **Mid** | | | | **High** | | | |
| **n=** | 39,617 | 33,458 | | | | 32,702 | | | |
|  | **%** | **%** | **aRR** | **(95% CI)** | p | **%** | **aRR** | **(95% CI)** | p |
| **Hospitalization** | 18.8 | 21.3 | 1.13 | (1.10–1.17) | <0.001 | 24.0 | 1.28 | (1.23–1.32) | <0.001 |
| Inpatient | 13.8 | 15.2 | 1.10 | (1.06–1.15) | <0.001 | 16.8 | 1.22 | (1.17–1.28) | <0.001 |
| Observation | 9.0 | 10.3 | 1.15 | (1.09–1.21) | <0.001 | 11.9 | 1.33 | (1.25–1.40) | <0.001 |
| **Cardiac Testing 7 days** | 11.2 | 12.6 | 1.13 | (1.07–1.19) | <0.001 | 13.7 | 1.23 | (1.17–1.28) | <0.001 |
| Stress ECG | 9.1 | 10.3 | 1.14 | (1.08–1.20) | <0.001 | 11.2 | 1.23 | (1.17–1.30) | <0.001 |
| Stress MIBI | 6.3 | 7.2 | 1.14 | (1.07–1.21) | <0.001 | 8.0 | 1.26 | (1.18–1.35) | <0.001 |
| Stress Echocardiogram | 2.5 | 2.9 | 1.17 | (1.04–1.29) | 0.009 | 3.0 | 1.21 | (1.07–1.34) | 0.003 |
| Stress PET | 0.18 | 0.18 | 0.99 | (0.61–1.36) | 0.939 | 0.18 | 0.99 | (0.72–1.26) | 0.959 |
| CCTA | 0.59 | 0.70 | 1.19 | (0.96–1.42) | 0.101 | 0.79 | 1.34 | (1.01–1.67) | 0.042 |
| **Cardiac Testing 30 days** | 16.1 | 17.8 | 1.10 | (1.06–1.15) | <0.001 | 18.7 | 1.16 | (1.12–1.21) | <0.001 |
| Stress ECG | 13.1 | 14.6 | 1.11 | (1.07–1.16) | <0.001 | 15.4 | 1.17 | (1.12–1.22) | <0.001 |
| Stress MIBI | 8.4 | 9.3 | 1.11 | (1.06–1.17) | <0.001 | 10.0 | 1.20 | (1.14–1.26) | <0.001 |
| Stress Echocardiogram | 3.7 | 4.2 | 1.13 | (1.03–1.22) | 0.011 | 4.2 | 1.14 | (1.04–1.24) | 0.005 |
| Stress PET | 0.23 | 0.26 | 1.13 | (0.75–1.51) | 0.500 | 0.24 | 1.02 | (0.79–1.26) | 0.854 |
| CCTA | 0.77 | 0.87 | 1.12 | (0.93–1.32) | 0.211 | 0.98 | 1.26 | (0.99–1.54) | 0.058 |
| **Invasive Care 30 days** |  |  |  |  |  |  |  |  |  |
| Angiogram | 7.2 | 7.8 | 1.09 | (1.03–1.14) | 0.002 | 8.3 | 1.15 | (1.09–1.20) | <0.001 |
| Coronary Intervention | 4.0 | 4.3 | 1.08 | (1.00–1.16) | 0.048 | 4.5 | 1.14 | (1.06–1.23) | <0.001 |
| PCI | 3.3 | 3.5 | 1.07 | (0.99–1.16) | 0.091 | 3.7 | 1.13 | (1.04–1.22) | 0.004 |
| CABG | 0.74 | 0.85 | 1.15 | (0.96–1.34) | 0.111 | 0.93 | 1.26 | (1.06–1.47) | 0.013 |
| **Invasive Care 180 days** |  |  |  |  |  |  |  |  |  |
| Angiogram | 8.3 | 8.97 | 1.08 | (1.03–1.13) | 0.002 | 9.4 | 1.13 | (1.08–1.19) | <0.001 |
| Coronary Intervention | 4.42 | 4.67 | 1.06 | (0.98–1.13) | 0.139 | 4.93 | 1.11 | (1.04–1.19) | 0.003 |
| PCI | 3.66 | 3.85 | 1.05 | (0.97–1.13) | 0.210 | 4.04 | 1.1 | (1.02–1.18) | 0.014 |
| CABG | 0.86 | 0.97 | 1.12 | (0.94–1.30) | 0.195 | 1.03 | 1.19 | (1.01–1.37) | 0.043 |
| **AMI Admissions** |  |  |  |  |  |  |  |  |  |
| Index | 2.8 | 3.1 | 1.1 | (1.00–1.20) | 0.048 | 3.3 | 1.16 | (1.06–1.25) | 0.001 |
| 7 day | 0.46 | 0.46 | 1.01 | (0.80–1.22) | 0.903 | 0.46 | 1.00 | (0.79–1.22) | 0.971 |
| 30 day | 0.58 | 0.57 | 0.98 | (0.80–1.16) | 0.843 | 0.59 | 1.02 | (0.83–1.21) | 0.851 |
| 180 days | 0.87 | 0.85 | 0.97 | (0.83–1.12) | 0.731 | 0.85 | 0.97 | (0.82–1.12) | 0.712 |
| **Abbreviations**: aRR, Adjusted risk ratio; CI, confidence interval; AMI, acute myocardial infarction; ECG, electrocardiogram; MIBI, sestamibi scan; PET, positron emission tomography; CCTA, cardiac computed tomography angiography; PCI, percutaneous coronary intervention; CABG, coronary artery bypass graft; ED, emergency department. | | | | | | | | | |
| **Note**: Clinician tendency was defined by the first 5 visits to each included clinician. Each included clinician were required to have ≥8 visits, with each ED having 2 such clinicians to be included. The denominator of the adjusted rates included all index chest pain ED visits that were the 6^th^ or later visits in the database for each clinician. The aRR were calculated in comparison to the adjusted rates of bottom-tercile clinicians. | | | | | | | | | |

| **Table S6. Adjusted Rates of Downstream Utilization and AMI Hospitalization by Clinician Admitting Tendency Defined by All ED Visits.** | | | | | | | | | |
| --- | --- | --- | --- | --- | --- | --- | --- | --- | --- |
| **Clinician Admitting Tendency** | Low | Mid | | | | High | | | |
| **n=** | 53,757 | 57,246 | | | | 48,000 | | | |
|  | **%** | **%** | **aRR** | **(95% CI)** | p | **%** | **aRR** | **(95% CI)** | p |
| **Hospitalization** | 9.5 | 20.5 | 2.17 | (2.08–2.25) | <0.001 | 33.7 | 3.57 | (3.40–3.73) | <0.001 |
| Inpatient | 7.2 | 15.0 | 2.09 | (1.99–2.20) | <0.001 | 22.6 | 3.16 | (2.97–3.34) | <0.001 |
| Observation | 4.4 | 9.7 | 2.23 | (2.09–2.37) | <0.001 | 17.2 | 3.95 | (3.64–4.25) | <0.001 |
| **Cardiac Testing 7 days** | 8.0 | 11.8 | 1.48 | (1.41–1.55) | <0.001 | 16.5 | 2.07 | (1.96–2.18) | <0.001 |
| Stress ECG | 6.6 | 9.8 | 1.47 | (1.40–1.54) | <0.001 | 13.6 | 2.05 | (1.93–2.17) | <0.001 |
| Stress MIBI | 4.2 | 7.1 | 1.7 | (1.60–1.80) | <0.001 | 10.3 | 2.48 | (2.31–2.64) | <0.001 |
| Stress Echocardiogram | 1.9 | 2.4 | 1.27 | (1.12–1.42) | <0.001 | 3.1 | 1.67 | (1.44–1.89) | <0.001 |
| Stress PET | 0.1 | 0.13 | 1.32 | (0.73–1.91) | 0.282 | 0.17 | 1.75 | (0.77–2.73) | 0.132 |
| CCTA | 0.29 | 0.59 | 2.03 | (1.34–2.71) | 0.003 | 0.9 | 3.09 | (1.68–4.49) | 0.004 |
| **Cardiac Testing 30 days** | 13.2 | 16.9 | 1.28 | (1.23–1.32) | <0.001 | 21.1 | 1.60 | (1.54–1.66) | <0.001 |
| Stress ECG | 11.1 | 14.0 | 1.26 | (1.22–1.31) | <0.001 | 17.6 | 1.59 | (1.52–1.65) | <0.001 |
| Stress MIBI | 6.4 | 9.2 | 1.43 | (1.37–1.50) | <0.001 | 12.3 | 1.91 | (1.81–2.00) | <0.001 |
| Stress Echocardiogram | 3.1 | 3.6 | 1.17 | (1.07–1.26) | <0.001 | 4.2 | 1.34 | (1.22–1.46) | <0.001 |
| Stress PET | 0.16 | 0.19 | 1.15 | (0.76–1.54) | 0.452 | 0.23 | 1.42 | (0.87–1.98) | 0.137 |
| CCTA | 0.43 | 0.79 | 1.81 | (1.36–2.26) | <0.001 | 1.04 | 2.39 | (1.56–3.21) | <0.001 |
| **Invasive Care 30 days** |  |  |  |  |  |  |  |  |  |
| Angiogram | 4.3 | 7.7 | 1.77 | (1.68–1.86) | <0.001 | 10.6 | 2.43 | (2.30–2.57) | <0.001 |
| Coronary Intervention | 2.2 | 4.3 | 1.92 | (1.77–2.06) | <0.001 | 5.9 | 2.64 | (2.43–2.85) | <0.001 |
| PCI | 2.2 | 3.9 | 1.78 | (1.65–1.91) | <0.001 | 5.2 | 2.38 | (2.20–2.56) | <0.001 |
| CABG | 0.54 | 0.95 | 1.76 | (1.51–2.02) | <0.001 | 1.31 | 2.42 | (2.07–2.78) | <0.001 |
| **Invasive Care 180 days** |  |  |  |  |  |  |  |  |  |
| Angiogram | 5.4 | 8.7 | 1.61 | (1.53–1.68) | <0.001 | 11.7 | 2.15 | (2.05–2.25) | <0.001 |
| Coronary Intervention | 2.6 | 4.7 | 1.77 | (1.65–1.89) | <0.001 | 6.3 | 2.40 | (2.23–2.57) | <0.001 |
| PCI | 2.2 | 3.9 | 1.78 | (1.65–1.91) | <0.001 | 5.2 | 2.38 | (2.20–2.56) | <0.001 |
| CABG | 0.54 | 0.95 | 1.76 | (1.51–2.02) | <0.001 | 1.31 | 2.42 | (2.07–2.78) | <0.001 |
| **AMI Admissions** | |  |  |  |  |  |  |  |  |
| Index | 1.5 | 3.2 | 2.04 | (1.86–2.22) | <0.001 | 4.3 | 2.79 | (2.53–3.05) | <0.001 |
| 7 day | 0.30 | 0.49 | 1.63 | (1.29–1.96) | <0.001 | 0.62 | 2.05 | (1.62–2.47) | <0.001 |
| 30 day | 0.39 | 0.62 | 1.60 | (1.32–1.88) | <0.001 | 0.74 | 1.90 | (1.56–2.25) | <0.001 |
| 180 days | 0.62 | 0.90 | 1.46 | (1.26–1.67) | <0.001 | 1.06 | 1.71 | (1.46–1.96) | <0.001 |
| **Abbreviations**: aRR, Adjusted risk ratio; CI, confidence interval; AMI, acute myocardial infarction; ECG, electrocardiogram; MIBI, sestamibi scan; PET, positron emission tomography; CCTA, cardiac computed tomography angiography; PCI, percutaneous coronary intervention; CABG, coronary artery bypass graft; ED, emergency department. | | | | | | | | | |
| **Note**: Clinician tendency was defined by examining hospitalization rates of all visits to each clinician (instead of only the first three). The denominator of the adjusted rates included all index chest pain ED visits that were the 4^th^ or later visits in the database for each clinician. The aRR were calculated in comparison to the adjusted rates of bottom-tercile clinicians. | | | | | | | | | |
